# The Balancing Role of Distribution Speed against Varying Efficacy Levels of COVID-19 Vaccines under Variants

**DOI:** 10.1101/2021.04.09.21255217

**Authors:** Daniel Kim, Pınar Keskinocak, Pelin Pekgün, Inci Yildirim

## Abstract

Mutations in SARS-CoV-2 raised concerns about diminishing vaccine effectiveness against COVID-19 caused by particular variants. Even with high initial efficacy, if a vaccine’s efficacy drops significantly against variants, or if it cannot be distributed quickly, it is uncertain whether the vaccine can provide better health outcomes than other vaccines. Hence, we evaluated the trade-offs between the *speed of distribution* vs. *efficacy against infection* of multiple vaccines when variants emerge by utilizing a Susceptible-Infected-Recovered-Deceased (SIR-D) model and assessing the level of *infection attack rate* (IAR). Our results show that speed is a key factor to a successful immunization strategy to control the COVID-19 pandemic even when the emerging variants may reduce the efficacy of a vaccine. Due to supply-chain challenges, the accessibility and distribution of the vaccines have been hindered in many regions, especially in low-income countries, while the second or third wave of the pandemic has occurred due to the variants. Understanding the tradeoffs between speed and efficacy and distributing vaccines that are available as quickly as possible are crucial to eradicate the pandemic before new variants spread.

## INTRODUCTION

Since the initial reports of a cluster of pneumonia cases of unidentified origin in Wuhan, China, in December 2019, more than 265 million people around the world have been infected with the novel severe acute respiratory syndrome coronavirus 2 (SARS-CoV-2).^1^ Despite the development of effective vaccines in unprecedented speed, concerns have been raised on the potential reduction in efficacy of these vaccines against the new SARS-CoV-2 variants due to possible evasion from antibody recognition.^2,3^ In order to reach herd immunity, effective implementation of a vaccine with sufficient efficacy against the circulating dominant variants is essential. Subsequently, it becomes a trivial decision for policymakers and governments to favor a vaccine with high efficacy for distribution. However, if the vaccine cannot be dispensed quickly and/or if its efficacy drops significantly against the emerging variants compared to other vaccines, the question of which vaccine should be favored is no longer trivial. Hence, the goal of this study was to understand the tradeoffs between the speed of distribution vs. the change in the efficacy levels of vaccines against infection before and after the emergence of variants, which we referred to as “initial efficacy” and “final efficacy”, respectively, hereafter.

The slow speed of procurement and dissemination of the SARS-CoV-2 vaccines has been a continual challenge, particularly for low-income countries. Many low-income countries could not even procure the small amount of vaccines since the high-income countries had reserved large amounts.^4^ In addition, the vaccines, especially those with high efficacies, may encounter a number of administrative and supply-chain related challenges in the low-income countries. For instance, the mRNA vaccines for COVID-19 require ultra-cold storage and logistics, which are often not readily available or easy to acquire.^5,6^ Due to such challenges, as of December 2021, only 8% of people in low-income countries have received at least one dose of the vaccines, in contrast to 65% of people in high-income countries.^7,8^

The distributional challenges and delays lead to continuous infections, providing an opportunity to the variants of the virus to emerge, which has raised concerns regarding reduced efficacy of vaccines against emerging variants.^9^ As of December 2021, five concerning SARS-CoV-2 variants have been identified: B.1.1.7, B.1.351, P.1, B.1.617.2, and B.1.1.529.^10^ These have been classified as the variants of concern (VOC) because they have quickly become the globally dominant forms.^11-14^ The variant B.1.617.2 (delta), for instance, was classified as the variant of concern by the World Health Organization (WHO) on May 11^th^, 2021 and quickly became dominant in the United States in July 2021.^10,15^ These variants became more alarming as multiple studies showed that the effectiveness of various vaccines decreased against the variants.^16-19^

In this paper, we studied the trade-offs between vaccines’ efficacy levels, which were subject to reduction due to emerging variants and speed of vaccine distribution. We developed an extended Susceptible-Infected-Recovered-Deceased (SIR-D) simulation model and assessed the *infection attack rate* (IAR) under different times that the virus variants emerge. A number of studies used the extended SIR-D model with different complications to study the impact of public health interventions, including social distancing and vaccination.^20-24^ While some papers examined the trade-offs between vaccine efficacy and distribution speed like ours,^23,24^, to the best of our knowledge, this is the first paper to consider the impact of a change in vaccine efficacy due to the emergence of variants. Throughout this paper, we referred to vaccine distribution as the entire distribution process of a vaccine including delivery to the dispensation sites and administration to the population. The results of this study were aimed to guide decision-makers in vaccine ordering during a pandemic when there are multiple types of vaccines, facing reduced efficacies as variants emerge.

## METHODS

### Vaccine Capacity

To compare different vaccine types, we categorized the level of the vaccine efficacy into three ranges: “H” (High) if 90% or above, M (Moderate) if higher than 70% and lower than 90%, and L (Low) if lower than 70%. We assumed that the final efficacy was always lower than the initial efficacy. In our main simulation, we considered three initial efficacy levels (*H*_*i*_ = 95%, *M*_*i*_ = 75%, and *L*_*i*_ = 65%) and three final efficacy levels (*H* _*f*_ = 90%, *M*_*f*_ = 70%, and *L*_*f*_ = 60%). Consequently, we obtained six types of vaccines, defined by a particular initial and final efficacy, as summarized in Table 1. These modeling choices were motivated by recent studies on vaccine efficacy against variants.^25-27^

**Table 1:** Vaccine Efficacy

| Vaccine Type | Initial Efficacy | Final Efficacy |
| --- | --- | --- |
| $H_i H_f$ | 95% | 90% |
| $H_i M_f$ | 95% | 70% |
| $H_i L_f$ | 95% | 60% |
| $M_i M_f$ | 75% | 70% |
| $M_i L_f$ | 75% | 60% |
| $L_i L_f$ | 65% | 60% |

We assumed in our simulations that all vaccines required a single dose and an individual who received an *effective* vaccine became fully protected against the disease upon vaccination. In each simulation, only a single type of vaccine was administered, and the daily vaccine distribution capacity was kept constant at *λ* · *K*, where *K* represents base capacity and *λ* is a multiplier. We fixed the base capacity, *K*, at 500,000, motivated from the average number of vaccine recipients in each day in the United States from December 14, 2020 to March 2, 2021, and we set a range of 1.0 to 3.0 with increments of 0.2 for the capacity multiplier *λ* to represent the speed of distribution in our simulations.^28^

### Compartmental Epidemiological Model

In our study, we utilized an extended SIR-D (Susceptible-Infectious-Recovered-Deceased) compartmental model, which is a simplified mathematical model of infectious diseases. In this model, individuals are moving among compartments, and transitions between compartments are governed by ordinary differential equations given epidemiological and vaccine parameters. We implemented seven compartments: *Susceptible* (*S*), *Vaccinated with immunity* (*V*), *Vaccinated-susceptible* (*S*^*v*^), *Symptomatic-infected* (*I*_*S*_), *Asymptomatic-infected* (*I*_*A*_), *Recovered* (*R*), and *Deceased* (*D*). When *Susceptible* population received vaccines, they entered either the *Vaccinated with immunity* (*V*) compartment if the vaccine was effective, or the *Vaccinated-susceptible* (*S*^*v*^) compartment, otherwise. Both Susceptible and Vaccinated-susceptible populations transitioned to either the *Symptomatic-infected* (*I*_*S*_) or *Asymptomatic-infected* (*I*_*A*_) compartment, once they made infectious contacts with the infected population. Symptomatic-infectious population then moved to either the *Recovered* (*R*) or *Deceased* (*D*) compartment. We assumed that asymptomatic patients always recovered in our model. The transition diagram of the extended SIR-D model is depicted in Figure 1.

**Figure 1:**
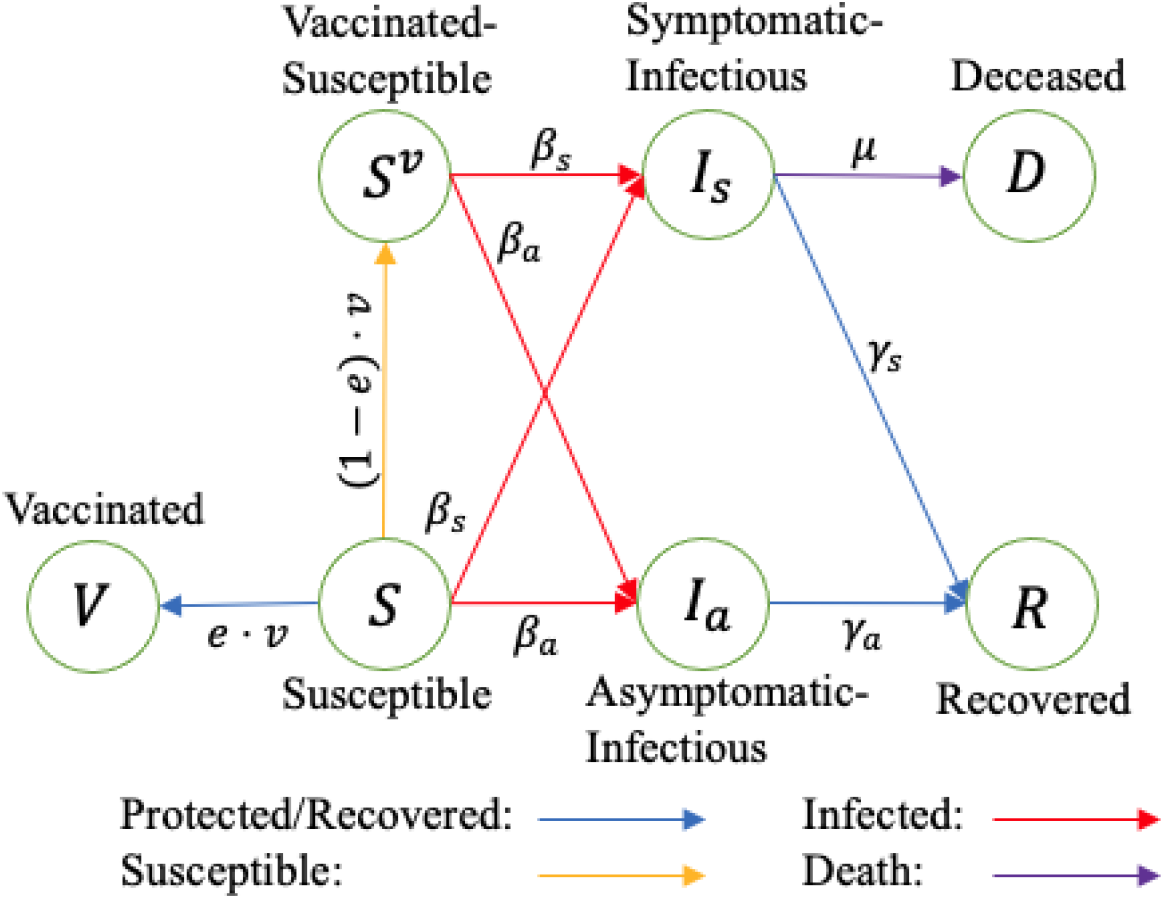
Transition diagram of the extended SIR-D model, in which each move is dependent on the epidemiological and vaccine parameters; symptomatic and asymptomatic transmission rates, respectively; : symptomatic and asymptomatic recovery rates, respectively; : decease rate of a symptomatic patient; : efficacy of the vaccine; : ratio of the daily vaccine capacity to the size of the unvaccinated susceptible population (i.e.,)

We chose the epidemiological parameter values that govern the transitions in the compartment model based on the estimated SARS-CoV-2 characteristics in various studies. The infectious periods of a symptomatic patient and an asymptomatic patient were studied to be 14 days and 8 days, respectively.^29,30^ Therefore, we set the recovery rates of symptomatic patients () and asymptomatic patients () at 1/14 and 1/8, respectively. In addition, the CDC has developed COVID-19 Pandemic Planning Scenarios and provided parameter values to use in a mathematical model.^31^ Accordingly, we assumed that 60% of the susceptible population who made infectious contacts became symptomatic. In addition, we used 2.3 for the reproduction number (), which is often employed by epidemiologists to represent the infectivity of a disease. The symptomatic infection fatality rate (IFR-S) of COVID-19, the proportion of deaths among symptomatic infected individuals, was estimated as 1.3%.^32^ Given the reproduction number of 2.3 and the IFR-S, we set the symptomatic-transmission rate (*β*_*s*_) to be 0.22031 and the death rate (*µ*) to be 0.0032. We assumed that the asymptomatic-transmission rate (*β*_*a*_) was 75% of the symptomatic-transmission rate.^33^ We used R-software to run the simulations with a population size (*N*) of 330 million (approximate population of the United States). Since our main goal was to analyze the trade-offs between distribution speed and vaccine’s efficacy under variants, we started the simulation only after when the vaccine became available and initialized it such that around 28% of the population had already been infected. Thus, we set 2.90% of the population as symptomatic-infected, 1.15% as asymptomatic-infected, 24.44% as recovered, and 0.14% as deceased. These estimates were motivated by the *confirmed* cumulative cases and deaths as of December 14, 2020, the first day of vaccine distribution in the United States.^34^ However, we multiplied the number of *confirmed* infections by six and increased the number of *confirmed* deaths by 35% in line with the findings of Wu et al. (2020) and Noh and Danuser (2021), who reported that the number of COVID-19 confirmed cumulative cases was underestimated, and Kung et al. (2020), who showed the same for confirmed deaths.^35-37^ The initial values in other compartments were estimated using the epidemiological parameters defined previously. All parameters used in the extended SIR-D model are summarized in Table 2 and the non-linear system of ordinary differential equations (ODEs) is as follows:

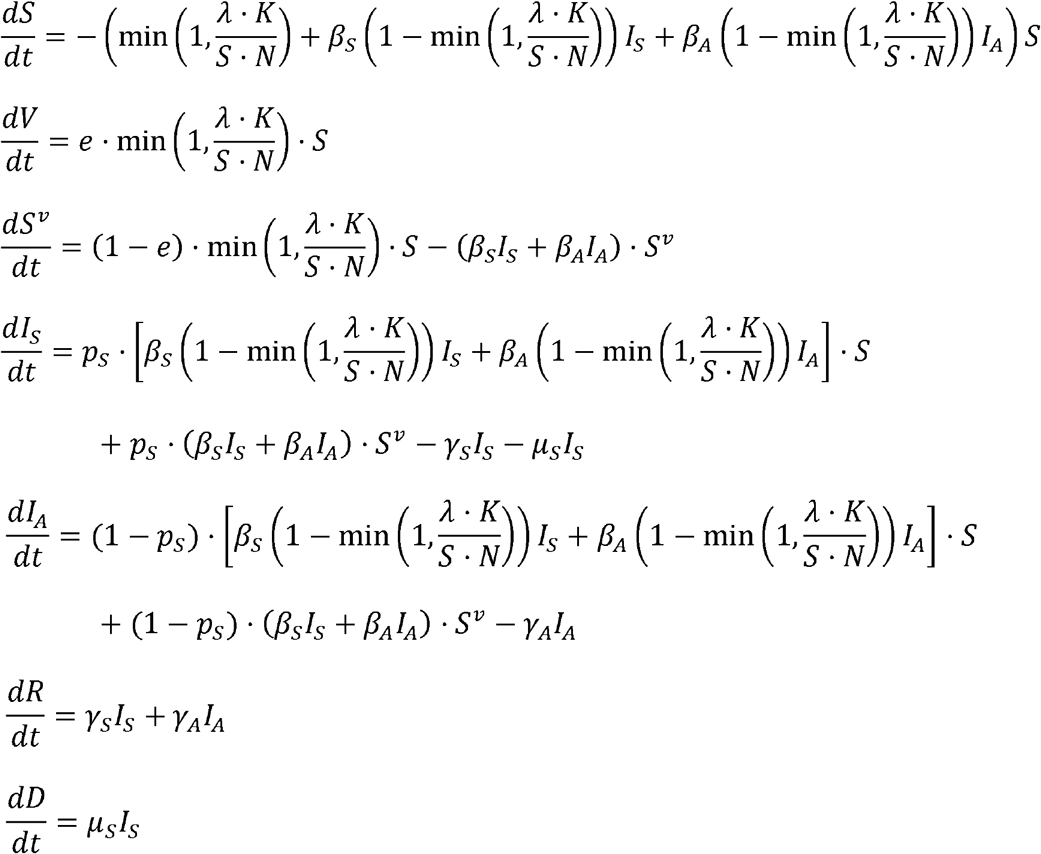

We ran the simulation on a one-year planning horizon under different *mutation times* (i.e., the time at which the emerging variants becomes dominant and cause a decrease in a vaccine’s efficacy) within the range of day 5 to day 40 with a discrete step size of 5 days, and different *capacity multipliers* (*λ*) within the range of 1.0 to 3.0 with a discrete step size of 0.2 to capture the vaccine distribution speed. We evaluated the impact of each vaccine type using IAR as the main health outcome.

**Table 2:** Parameters used in the extended SIR-D Model

| Parameter | Description | Value |
| --- | --- | --- |
| $\beta_S, \beta_A$ | Transmission rate | 0.22031, 0.16523 |
| $\gamma_S, \gamma_A$ | Recovery rate | 1/14, 1/8 |
| $\mu_S$ | Death rate | 0.0032 |
| $N$ | Population size | 330 million |
| $p_S$ | Probability of developing symptoms | 0.6 |
| $e$ | Vaccine efficacy | See Table 1 |
| $K$ | Base capacity | 500,000 |
| $\lambda$ | Capacity multiplier | 1.0 to 3.0 with increments of 0.2 |

## RESULTS

We first ran our simulation without the presence of any vaccines, which gave an estimated IAR of approximately 81% and IFR-S of 2%. The daily infection peak (i.e., the highest percentage of the population who get newly infected on a single day) occurred on day 24, at which 1% of the population got newly infected.

Table 3 shows the estimated IAR under different capacity multipliers (*λ*) when the mutation times are day 5 and day 25. We report the full results with different mutation times in Supplemental Materials. When the mutation time is day 5, the level of IAR is between 60% and 65% when the capacity multiplier is 3.0 (1.5M doses/day) and between 72% and 75% when the capacity multiplier is 1.0 (0.5M doses/day), depending on the selected vaccine type. When the mutation time is day 25, the level of IAR is smaller compared to when the mutation time is day 5, and the decrease in IAR due to the increase in capacity multiplier is larger.

**Table 3:**
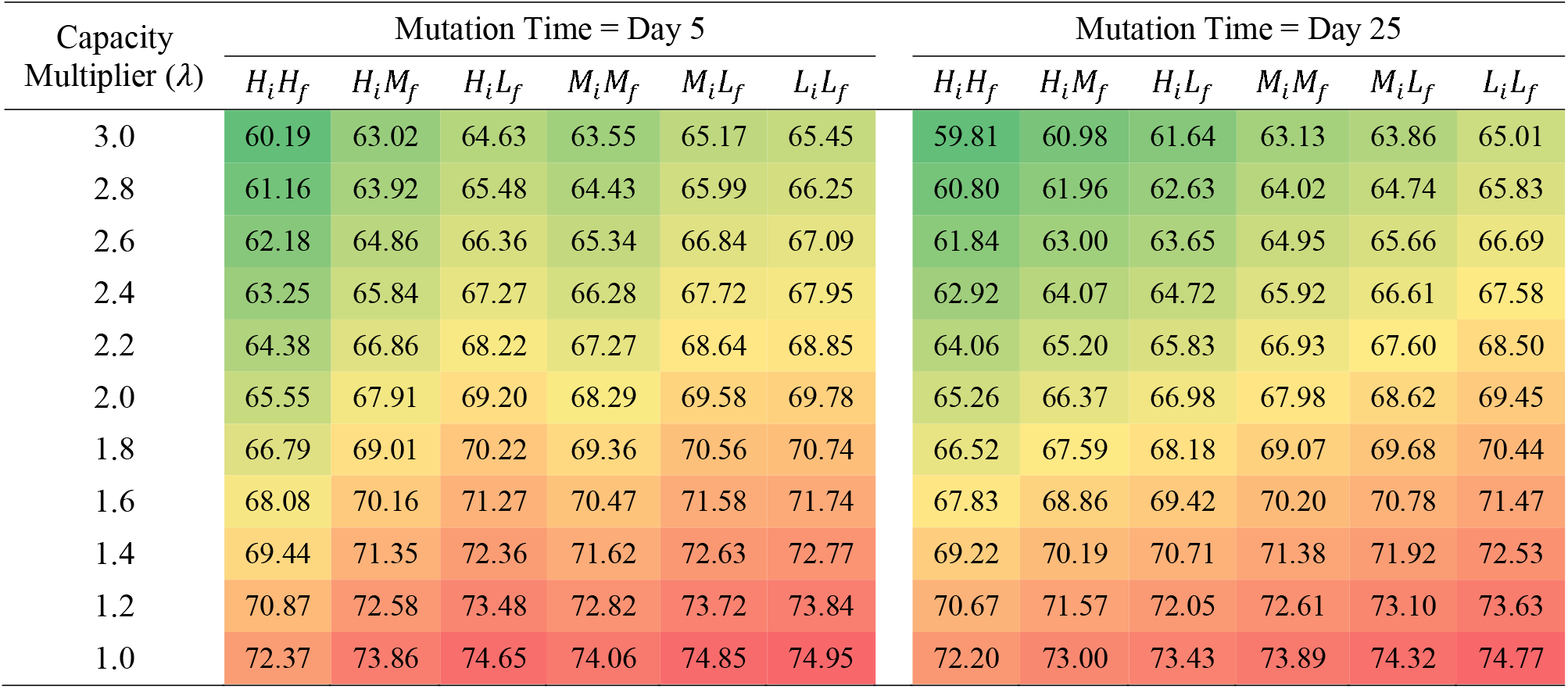
Infection attack rate (%) under different capacity multipliers and vaccine types when mutation time is day 5 and day 25

| Capacity Multiplier ($\lambda$) | Mutation Time = Day 5 | | | | | | Mutation Time = Day 25 | | | | | |
| --- | --- | --- | --- | --- | --- | --- | --- | --- | --- | --- | --- | --- |
| | $H_i H_f$ | $H_i M_f$ | $H_i L_f$ | $M_i M_f$ | $M_i L_f$ | $L_i L_f$ | $H_i H_f$ | $H_i M_f$ | $H_i L_f$ | $M_i M_f$ | $M_i L_f$ | $L_i L_f$ |
| 3.0 | 60.19 | 63.02 | 64.63 | 63.55 | 65.17 | 65.45 | 59.81 | 60.98 | 61.64 | 63.13 | 63.86 | 65.01 |
| 2.8 | 61.16 | 63.92 | 65.48 | 64.43 | 65.99 | 66.25 | 60.80 | 61.96 | 62.63 | 64.02 | 64.74 | 65.83 |
| 2.6 | 62.18 | 64.86 | 66.36 | 65.34 | 66.84 | 67.09 | 61.84 | 63.00 | 63.65 | 64.95 | 65.66 | 66.69 |
| 2.4 | 63.25 | 65.84 | 67.27 | 66.28 | 67.72 | 67.95 | 62.92 | 64.07 | 64.72 | 65.92 | 66.61 | 67.58 |
| 2.2 | 64.38 | 66.86 | 68.22 | 67.27 | 68.64 | 68.85 | 64.06 | 65.20 | 65.83 | 66.93 | 67.60 | 68.50 |
| 2.0 | 65.55 | 67.91 | 69.20 | 68.29 | 69.58 | 69.78 | 65.26 | 66.37 | 66.98 | 67.98 | 68.62 | 69.45 |
| 1.8 | 66.79 | 69.01 | 70.22 | 69.36 | 70.56 | 70.74 | 66.52 | 67.59 | 68.18 | 69.07 | 69.68 | 70.44 |
| 1.6 | 68.08 | 70.16 | 71.27 | 70.47 | 71.58 | 71.74 | 67.83 | 68.86 | 69.42 | 70.20 | 70.78 | 71.47 |
| 1.4 | 69.44 | 71.35 | 72.36 | 71.62 | 72.63 | 72.77 | 69.22 | 70.19 | 70.71 | 71.38 | 71.92 | 72.53 |
| 1.2 | 70.87 | 72.58 | 73.48 | 72.82 | 73.72 | 73.84 | 70.67 | 71.57 | 72.05 | 72.61 | 73.10 | 73.63 |
| 1.0 | 72.37 | 73.86 | 74.65 | 74.06 | 74.85 | 74.95 | 72.20 | 73.00 | 73.43 | 73.89 | 74.32 | 74.77 |

Figure 2 and Figure 3 present the contour plot and the two-dimensional plot of the IAR in Table 3, respectively. Figure 2 shows that even vaccine-*L*_*i*_*L*_*f*_ can achieve a lower IAR than vaccine-*H*_*i*_*H*_*f*_ if the capacity multiplier of vaccine-*L*_*i*_*L*_*f*_ is high compared to that of vaccine-*H*_*i*_*H*_*f*_. For instance, when the capacity multiplier of vaccine-*H*_*i*_*H*_*f*_ is 1.0 and the mutation time is day 5, 72.3% of the population is infected. However, if the capacity multiplier of vaccine-*L*_*i*_*L*_*f*_ is 1.48 (or higher), 72.2% (or less) of the population is infected. We present the minimum required capacity multiplier of all vaccine types to achieve a lower IAR than vaccine-*H*_*i*_*H*_*f*_ with the capacity multiplier of 1.0 and 1.6 under different mutation times in Supplemental Materials.

**Figure 2:**
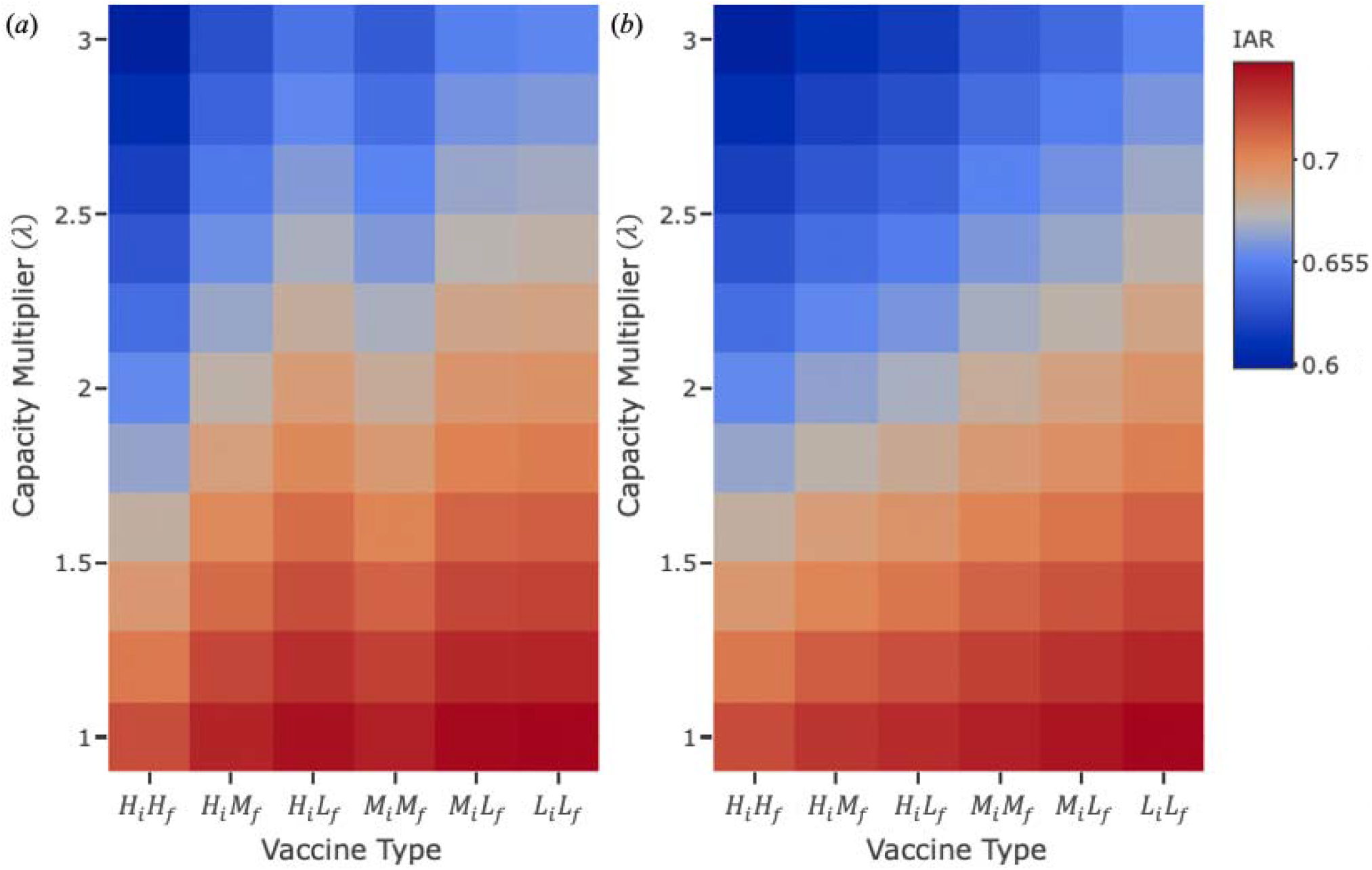
Graphical representation of infection attack rate under different capacity multipliers with different vaccine types when mutation time is (a) day 5 and (b) day 25

**Figure 3:**
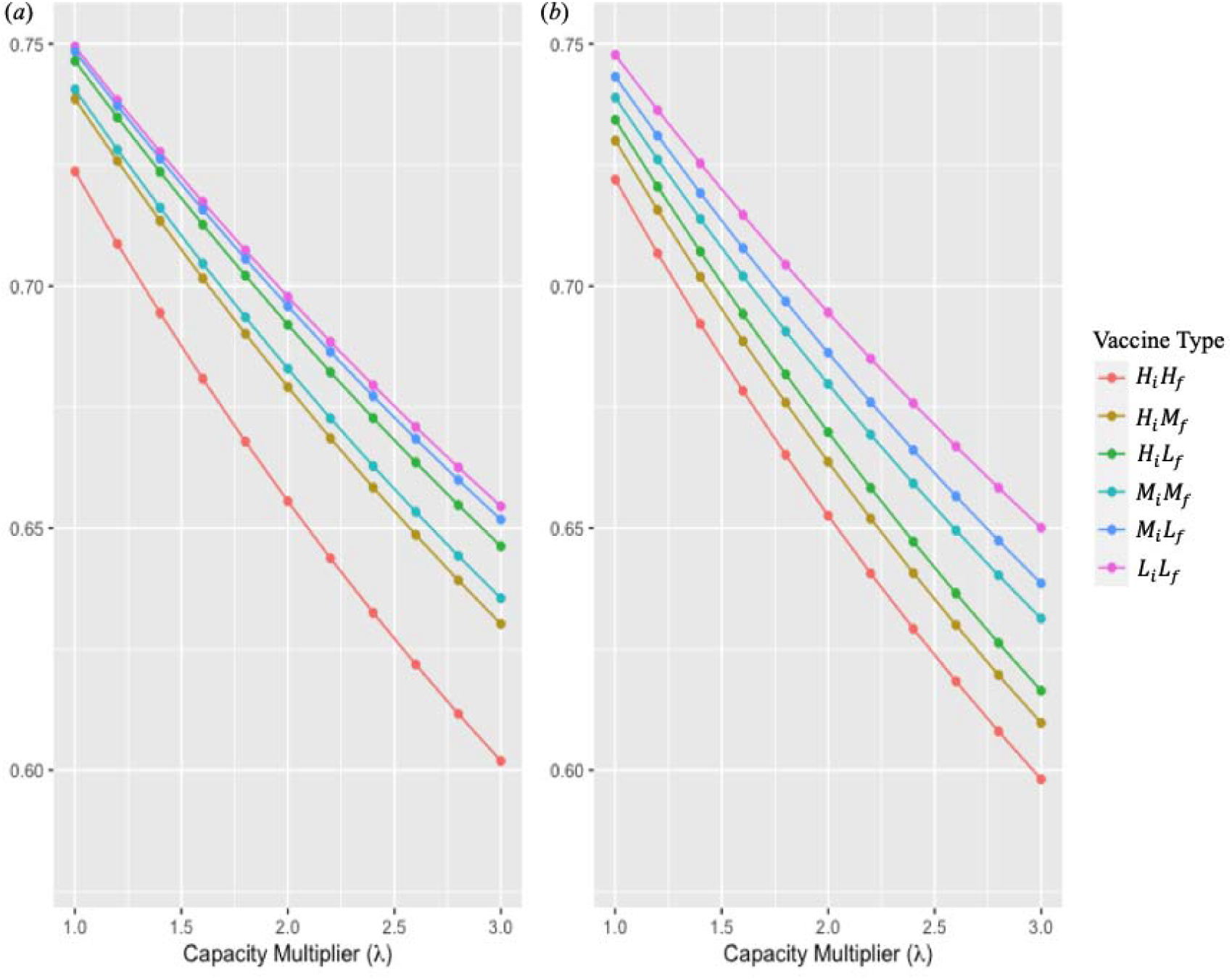
Two-dimensional plot of infection attack rate under different capacity multipliers with different vaccine types when mutation time is (a) day 5 and (b) day 25

Figure 4 compares the daily new infections from day 5 to day 50 with vaccine-*H*_*i*_*H*_*f*_ vs. *M*_*i*_*M*_*f*_ vaccine In this figure, the mutation time is early (on day 5) and comes before the daily infection peak, and thus, vaccine-*M*_*i*_*M*_*f*_ results in a better IAR than vaccine-*H*_*i*_*H*_*f*_ for all capacity multipliers. Specifically, after the peak of daily new infections is reached for each vaccine, the number of daily infections drops at a faster rate when vaccine-*M*_*i*_*M*_*f*_ is administered. On the other hand, when the mutation time comes after the daily infection peak (not shown in the figure), vaccine-*M*_*i*_*M*_*f*_ achieves a lower IAR than vaccine-*M*_*i*_*M*_*f*_ for all capacity multipliers.

**Figure 4:**
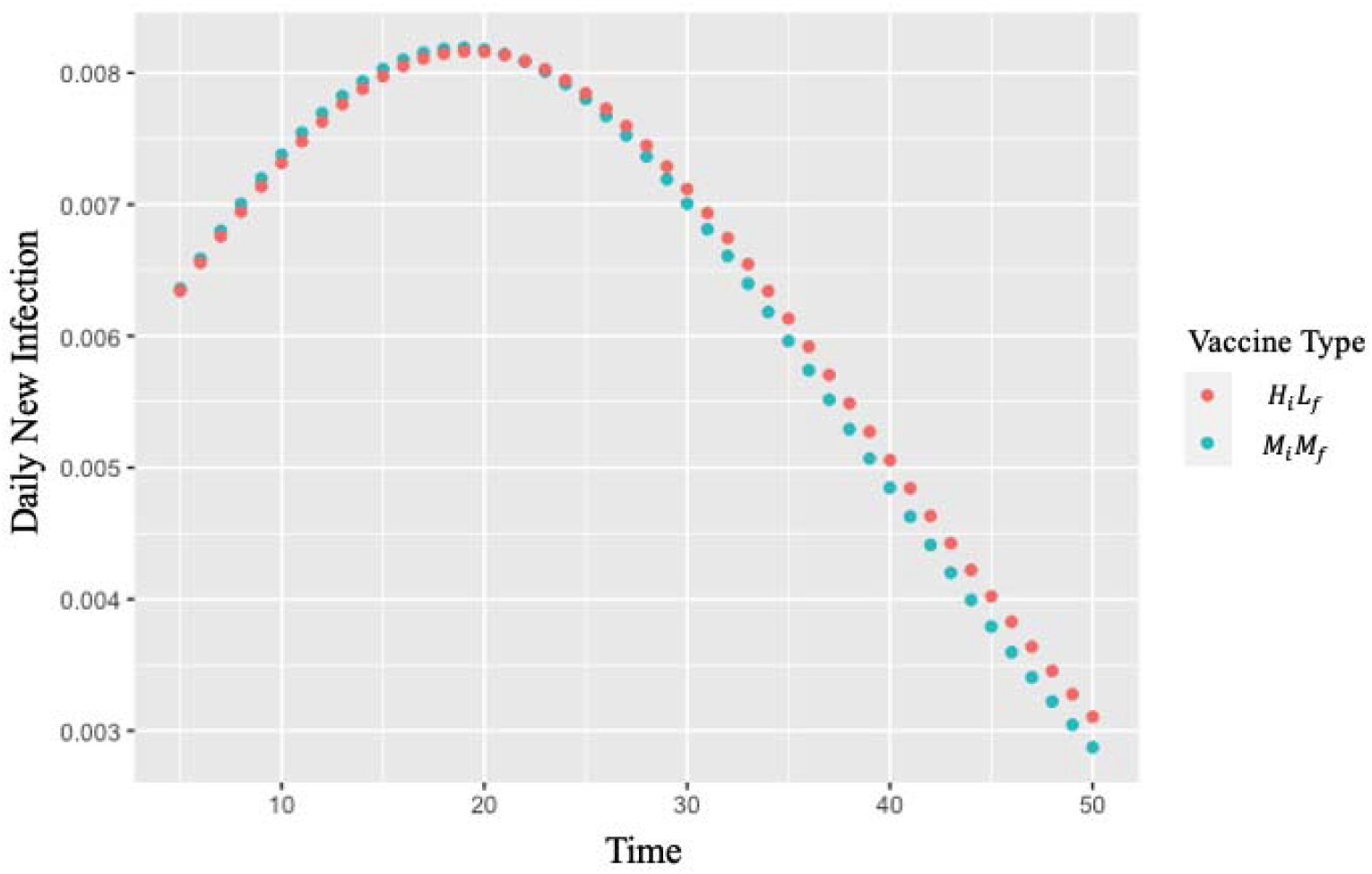
Daily new infections from day 5 to day 50 with vaccine- and vaccine- when the capacity multiplier (is 3 and the mutation time is day 5

## DISCUSSION

In this study, we utilized an extended SIR-D model to simulate the trajectory of an infectious disease under the intervention of different kinds of vaccines of which efficacies decrease against the variants of the disease and different mutation times. We have demonstrated that the speed of the vaccine distribution is a key factor to achieve low IAR levels, even though the vaccine may have high efficacy both before and after the variants emerge.

Our study showed that a vaccine with low initial and final efficacy levels (vaccine-) could achieve a lower IAR than a vaccine with high initial and final efficacy levels (vaccine-) if the former can be distributed more quickly than the latter, regardless of the mutation time. In our simulation, when the mutation time was day 25 and the capacity multiplier of vaccine- was 1.0 (0.5M doses/day), 72.2% of the population got infected. If the capacity multiplier of vaccine-*L*_*i*_*L*_*f*_ was 1.47 (0.73M doses/day or higher), 72.1% (or less) of the population was infected (Table 3). Even for vaccines with the same initial efficacy, such as vaccine-*H*_*i*_*H*_*f*_ and vaccine-*H*_*i*_*H*_*f*_, this result remained robust—vaccine-*H*_*i*_*H*_*f*_ with a capacity multiplier of 1.18 (0.59M doses/day) or higher achieved a lower IAR than vaccine-*H*_*i*_*H*_*f*_ with a capacity multiplier of 1.0 (0.5M doses/day). Since the start of the COVID-19 vaccination process, the speed of the vaccine distribution has been slow due to numerous reasons, including limited and uncertain vaccine supply and various logistics and storage challenges. Despite the continuing effort of increasing production capacities, vaccine manufacturers, especially those who produce mRNA vaccines with new technology, have been struggling to secure sufficient supply of vaccine ingredients, storage containers, and more, due to the demand from billions of people around the world.^38^ In addition, mRNA vaccines need to be stored in ultra-cold freezers under specific expiration dates, although many communities, especially in the low-income countries, lack or cannot afford such infrastructure, leading to a limited number of administration sites. Besides the mRNA vaccines, numerous vaccines that require the same level of the resources that the seasonal flu vaccine consumes have been developed and administered throughout the world. These vaccines may prevent vaccine wastage, enable efficient production and distribution using the existing vaccine supply chain, and facilitate a faster rate of vaccination.^39^ Hence, despite having lower efficacy than mRNA vaccines, those vaccines that have the potential for faster distribution may be more beneficial.

Increasing the doses distributed per day, i.e., the capacity multiplier (*λ*), of any vaccine type is always beneficial as it reduces the size of the susceptible population and can, eventually, achieve herd immunity. Our study showed that the level of IAR always decreased when the capacity multiplier was higher, with the largest impact observed for vaccine-*H*_*i*_*H*_*f*_. In particular, when we compared the impact of changing the capacity multiplier from 1.0 to 3.0 for vaccine-*H*_*i*_*H*_*f*_, vaccine-*M*_*i*_*M*_*f*_, and vaccine-*L*_*i*_*L*_*f*_, IAR changed from 72.20% to 59.81% for *H*_*i*_*H*_*f*_, from 73.89% to 63.13% for, and from 74.77% to 65.01% for, when the difference between the initial and final efficacy was 5% for all the vaccine types and the mutation time was day 25. In addition, if vaccine-*H*_*i*_*H*_*f*_ could be distributed at a faster rate, the minimum required capacity multiplier (*λ*) of vaccine-*L*_*i*_*L*_*f*_ to achieve a lower IAR than vaccine-*H*_*i*_*H*_*f*_was even larger. For example, when the mutation time was day 25, the capacity multiplier of vaccine-*L*_*i*_*L*_*f*_ needed to be at least 1.47 to achieve a lower IAR than vaccine-*H*_*i*_*H*_*f*_ with the capacity multiplier of 1.0. On the other hand, when the vaccine-*H*_*i*_*H*_*f*_’s capacity multiplier was 1.6, the capacity multiplier of vaccine-*L*_*i*_*L*_*f*_ needed to be at least 2.35. Thus, even though the difference in the capacity multiplier of vaccine-*H*_*i*_*H*_*f*_ was only 0.60, that of vaccine-*L*_*i*_*L*_*f*_ was 0.88. However, increasing the capacity multiplier, i.e., the speed of distribution, for vaccine-*H*_*i*_*H*_*f*_, which represents mRNA vaccines in our model, may be much more challenging than that for vaccine-*L*_*i*_*L*_*f*_, as described above, including economic burden, complications in vaccination programs, and unsophisticated infrastructure. In such cases, it may be more beneficial to allocate resources towards distributing a lower efficacy vaccine at a faster rate as our study shows.

Forecasting the time when the peak infections occur and when the variants emerge is also critical to choosing which type of vaccine to distribute for maximizing public health benefits. If the mutation time comes after the daily infection peak, a vaccine with a higher initial efficacy always achieves a lower IAR than a vaccine with lower initial efficacy. However, if the mutation time comes before the daily infection peak, the final efficacy level determines which vaccine type achieves a lower IAR under the same capacity multiplier. For example, when we compared vaccine-*H*_*i*_*H*_*f*_ and vaccine-*M*_*i*_*M*_*f*_ with a capacity multiplier of 1.0 for each, the daily infection peak occurred on day 22 for both vaccines. Then, the administration of vaccine-*M*_*i*_*M*_*f*_, which has an initial efficacy of 75% and final efficacy of 70%, resulted in an IAR of 74.06%, whereas the administration of vaccine-*H*_*i*_*H*_*f*_, which has an initial efficacy of 95% and final efficacy of 60%, resulted in an IAR of 74.65% (Figure 4). This is because an effective vaccination program achieves the highest reduction in the number of new infections *before* the daily infection peak. Afterwards, even with a higher initial efficacy, a vaccine with a lower final efficacy cannot reduce the size of the susceptible population as much as a vaccine with a lower initial efficacy and a higher final efficacy. Active genomic surveillance that studies the evolvement of the virus is critical to identify a new variant and study its influence on the spread of the disease and the vaccine efficacy.^40^ However the genomic surveillance has not received as much attention and the coverage is still low.^41,42^ Our results demonstrated that an expedited detection of the variants and their impact is vital to the choice of a vaccine to minimizes the IAR.

## Limitation

We acknowledge some limitations of this study. Our compartmental model provided insights on the trade-offs between speed and efficacy against emerging variants without incorporating the impact of other interventions. However, it can be extended to capture more realistic trajectory of SARS-CoV-2, including more compartments or time-dependent epidemiological parameters.^43,44^ In addition, we assumed that the population gains immunity as soon as they receive effective vaccines and that every type of vaccine requires a single dose. In practice, the majority of the authorized vaccines require two doses with three to four weeks apart application and it may take several days to gain immunity after vaccination. Moreover, we did not consider any type of non-pharmaceutical interventions. Depending on the number of people who conform to the interventions, such as social distancing and mask mandates, the probability of infectious contacts may vary over time.

## CONCLUSION

Overall, our results suggested that the administration of a vaccine with high efficacy against both the original strain and the variants may not always lead to a low number of cumulative infections if it cannot be distributed as quickly as other vaccine types with lower efficacies. Despite the vast efforts for worldwide vaccination, the vaccine distribution has been an ongoing challenge due to production shortages, economic constraints, and the lack of advanced supply-chain infrastructure, which is critical to distribute some of the high-efficacy vaccines. Due to these challenges, the accessibility and distribution of the vaccines have been hindered especially in many low- and middle-income countries.^45-47^ It is critical to distribute available vaccines as quickly as possible and vaccinate more people to reach herd immunity before new variants spread. Our study demonstrated that a vaccine with a relatively lower efficacy can achieve at least as good health outcomes as their higher efficacy counterparts, as long as it can be distributed more quickly. We hope that our study provides guidance to decision makers on the tradeoffs between speed and efficacy, highlighting the critical role of speed of vaccination during a pandemic as variants that decrease efficacy of vaccines emerge.

## Data Availability

No data is used or referred to in the manuscript.

## Supplemental Materials

**Supplemental Table 1:**
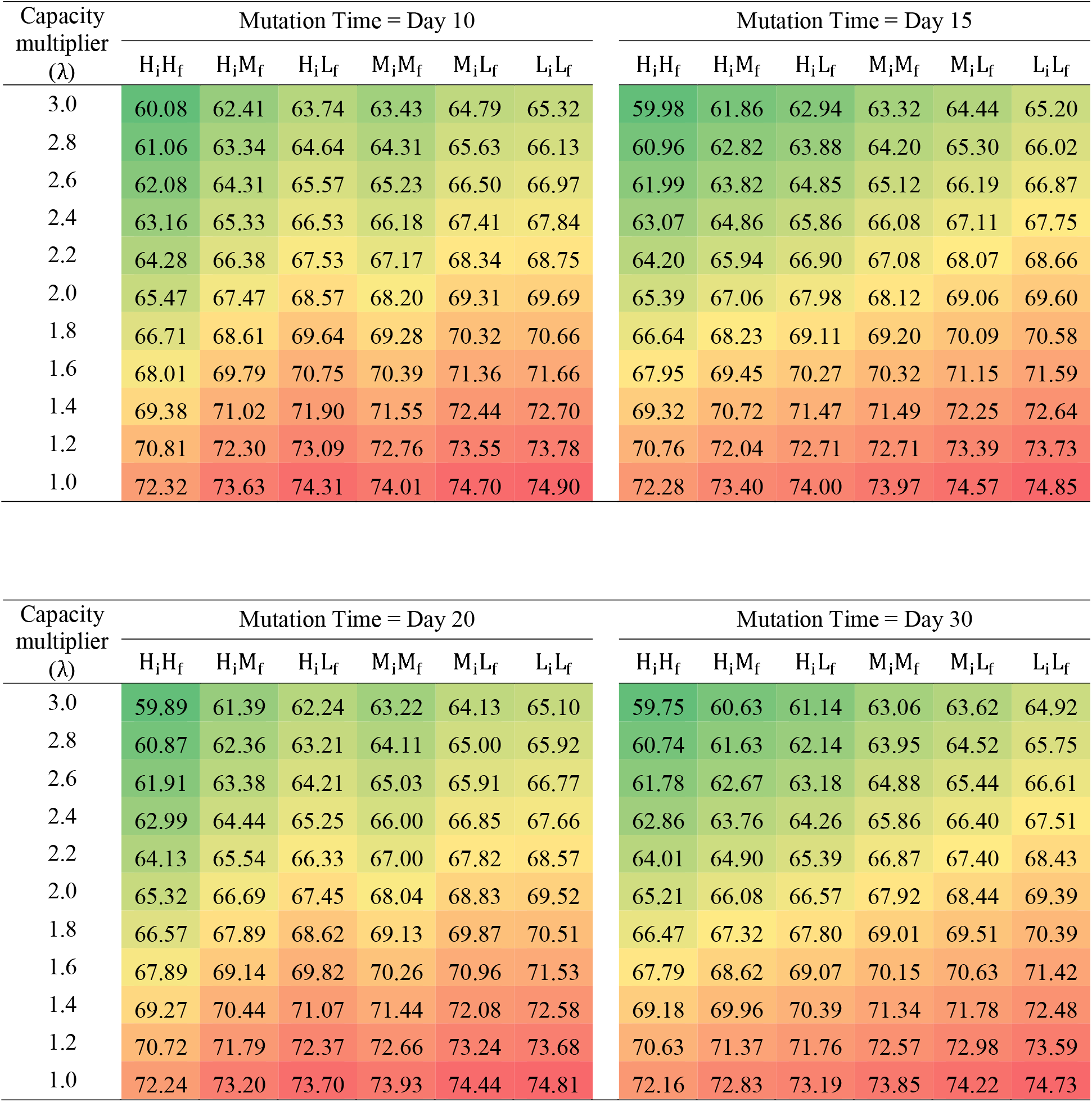
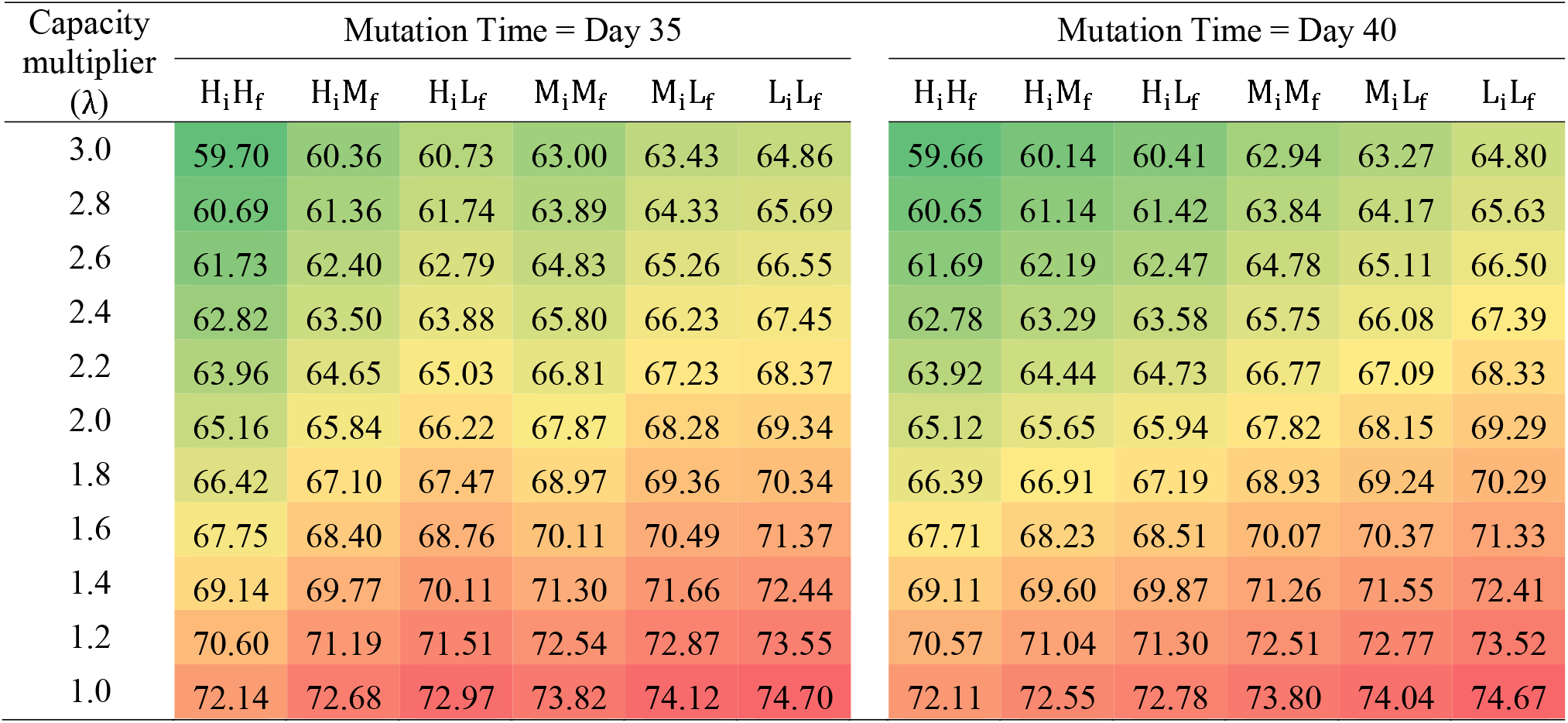
Infection attack rate (%) under different capacity multipliers and mutation times

**Supplemental Table 2:**
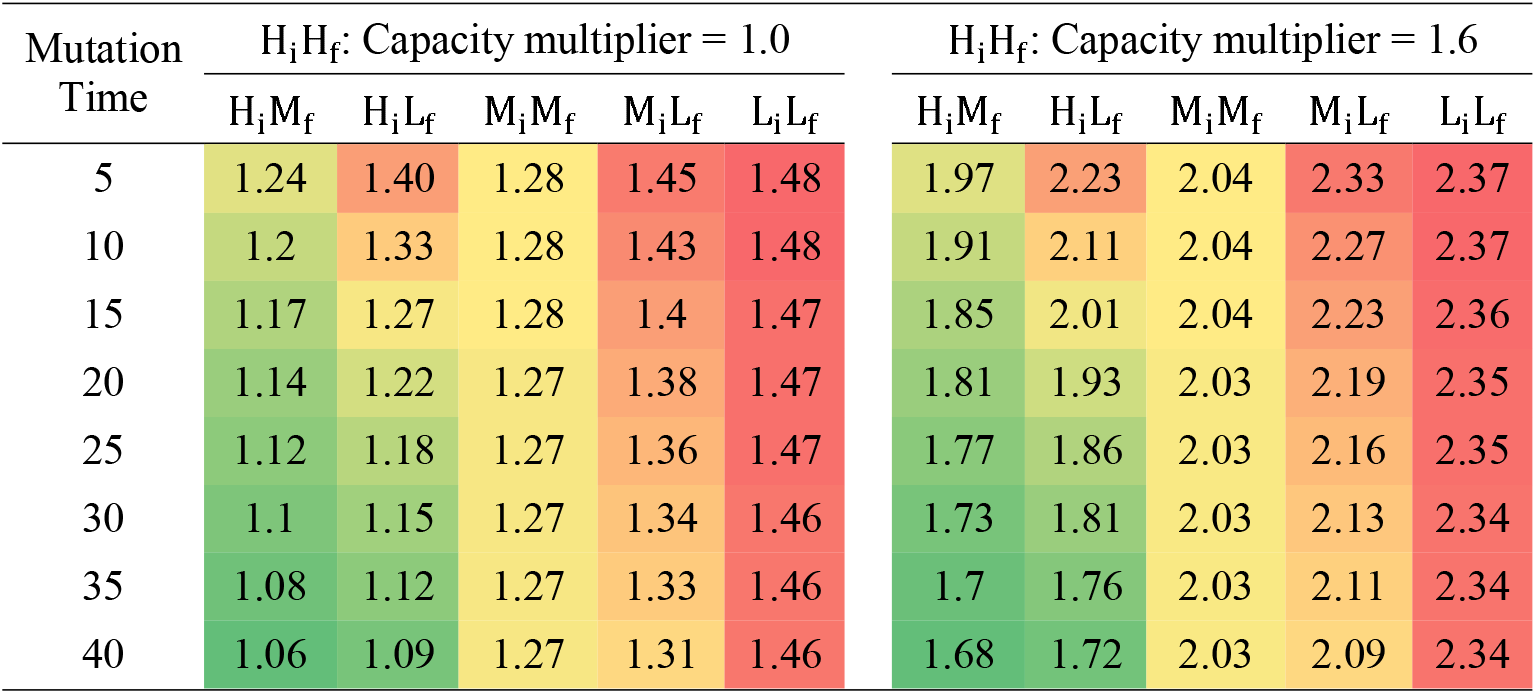
Minimum required capacity multiplier of each vaccine type under different mutation times to achieve a lower IAR than vaccine-H_i_H_f_ with the capacity multipliers of 1.0 and 1.6

## REFERENCES

1 World Health Organization. WHO Coronavirus Disease (COVID-19) Dashboard, <https://covid19.who.int> (2020).

2 Karim, S. S. A. & Karim, Q. A. Omicron SARS-CoV-2 variant: a new chapter in the COVID-19 pandemic. The Lancet (2021).

3 Sanders, R. W. & de Jong, M. D. Pandemic moves and countermoves: vaccines and viral variants. The Lancet (2021).

4 Choi, E. M. COVID-19 vaccines for low-and middle-income countries. Trans R Soc Trop Med Hyg 115, 447–456, doi:10.1093/trstmh/trab045 (2021).

5 Tregoning, J. S., Flight, K. E., Higham, S. L., Wang, Z. & Pierce, B. F. Progress of the COVID-19 vaccine effort: viruses, vaccines and variants versus efficacy, effectiveness and escape. Nature Reviews Immunology, 1–11 (2021).

6 Fischer, W. A., II, M. G., Bhagwanjee S. & Sevransky, J. Global burden of influenza: contributions from resource limited and low-income settings. Global heart 9, 325 (2014).

7 Hannah Ritchie et al. (Our World in Data, 2020).

8 Ramachandran, R., Ross, J. S. & Miller, J. E. Access to COVID-19 Vaccines in High-, Middle-, and Low-Income Countries Hosting Clinical Trials. JAMA Network Open 4, e2134233–e2134233 (2021).

9 Knight, K. & Radhakrishnan, A. Omicron Latest Reminder That Global Vaccine Equity Is Critical, 2021).

10 World Health Organization. Tracking SARS-CoV-2 variants, <https://www.who.int/en/activities/tracking-SARS-CoV-2-variants/> (2021).

11 Lauring, A. S. & Hodcroft, E. B. Genetic Variants of SARS-CoV-2—What Do They Mean? JAMA (2021).

12 Smith-Schoenwalder, C. CDC: Coronavirus Variant First Found in U.K. Now Dominant Strain in U.S., <https://www.usnews.com/news/health-news/articles/2021-04-07/cdc-coronavirus-variant-first-found-in-uk-now-dominant-strain-in-us> (2021).

13 Glatter, R. P.1 Variant, Dominant Strain in Brazil, Reported in New York, <https://www.forbes.com/sites/robertglatter/2021/03/21/p1-variant-dominant-strain-in-brazil-reported-in-new-york/?sh=449db2971883> (2021).

14 Adam, D. What scientists know about new, fast-spreading coronavirus variants, <https://doi.org/10.1038/d41586-021-01390-4> (2021).

15 Centers for Disease Control and Prevention. Variant Proportions, <https://covid.cdc.gov/covid-data-tracker/#variant-proportions> (2021).

16 Lopez Bernal, J. et al. Effectiveness of Covid-19 vaccines against the B. 1.617. 2 (Delta) variant. N Engl J Med, 585–594 (2021).

17 Abu-Raddad, L. J., Chemaitelly, H. & Butt, A. A. Effectiveness of the BNT162b2 Covid-19 Vaccine against the B.1.1.7 and B.1.351 Variants. New England Journal of Medicine 385, 187–189, doi:10.1056/NEJMc2104974 (2021).

18 Liu, C. et al. Reduced neutralization of SARS-CoV-2 B.1.617 by vaccine and convalescent serum. Cell, doi:https://doi.org/10.1016/j.cell.2021.06.020 (2021).

19 Wang, P. et al. Antibody resistance of SARS-CoV-2 variants B.1.351 and B.1.1.7. Nature 593, 130–135, doi:10.1038/s41586-021-03398-2 (2021).

20 Grimm, V., Mengel, F. & Schmidt, M. Extensions of the SEIR model for the analysis of tailored social distancing and tracing approaches to cope with COVID-19. Scientific Reports 11, 1–16 (2021).

21 Rădulescu, A., Williams, C. & Cavanagh, K. Management strategies in a SEIR-type model of COVID 19 community spread. Scientific reports 10, 1–16 (2020).

22 Usherwood, T., LaJoie, Z. & Srivastava, V. A model and predictions for COVID-19 considering population behavior and vaccination. Scientific Reports 11, 1–11 (2021).

23 Paltiel, A. D., Schwartz, J. L., Zheng, A. & Walensky, R. P. Clinical Outcomes Of A COVID-19 Vaccine: Implementation Over Efficacy: Study examines how definitions and thresholds of vaccine efficacy, coupled with different levels of implementation effectiveness and background epidemic severity, translate into outcomes. Health Affairs, 10.1377/hlthaff.2020.02054 (2021).

24 Paltiel, A. D., Zheng, A. & Schwartz, J. L. Speed versus efficacy: quantifying potential tradeoffs in COVID-19 vaccine deployment. Annals of internal medicine (2021).

25 Food and Drug Administration. Fact Sheet for Healthcare Providers Administering Vaccine (Vaccine Providers) Emergency Use Authorization (EUA) of the Janssen COVID-19 Vaccine to Prevent Coronavirus Disease 2019 (COVID-19), 2021).

26 Pfizer. Pfizer and BioNTech Confirm High Efficacy and No Serious Safety Concerns Through Up to Six Months Following Second Dose in Updated Topline Analysis of Landmark COVID-19 Vaccine Study, <https://www.pfizer.com/news/press-release/press-release-detail/pfizer-and-biontech-confirm-high-efficacy-and-no-serious> (2021).

27 Nanduri, S. et al. Effectiveness of Pfizer-BioNTech and Moderna vaccines in preventing SARS-CoV-2 infection among nursing home residents before and during widespread circulation of the SARS-CoV-2 B. 1.617. 2 (Delta) variant—National Healthcare Safety Network, March 1–August 1, 2021. Morbidity and Mortality Weekly Report 70, 1163 (2021).

28 Centers for Disease Control and Prevention. COVID-19 Vaccinations in the United States, <https://covid.cdc.gov/covid-data-tracker/#vaccinations> (2020).

29 You, C. et al. Estimation of the time-varying reproduction number of COVID-19 outbreak in China. International Journal of Hygiene and Environmental Health 228, 113555 (2020).

30 Byrne, A. W. et al. Inferred duration of infectious period of SARS-CoV-2: rapid scoping review and analysis of available evidence for asymptomatic and symptomatic COVID-19 cases. BMJ open 10, e039856 (2020).

31 Centers for Disease Control and Prevention. COVID-19 Pandemic Planning Scenarios, <https://www.cdc.gov/coronavirus/2019-ncov/hcp/planning-scenarios.html> (2020).

32 Basu, A. Estimating The Infection Fatality Rate Among Symptomatic COVID-19 Cases In The United States: Study estimates the COVID-19 infection fatality rate at the US county level. Health Affairs 39, 1229–1236 (2020).

33 Oran, D. P. & Topol, E. J. Prevalence of asymptomatic SARS-CoV-2 infection: a narrative review. Annals of internal medicine 173, 362–367 (2020).

34 Centers for Disease Control and Prevention. COVID Data Tracker, <https://covid.cdc.gov/covid-data-tracker/#datatracker-home> (2020).

35 Kung, S. et al. Underestimation of COVID-19 mortality during the pandemic. ERJ open research 7 (2021).

36 Wu, S. L. et al. Substantial underestimation of SARS-CoV-2 infection in the United States. Nature communications 11, 1–10 (2020).

37 Noh, J. & Danuser, G. Estimation of the fraction of COVID-19 infected people in US states and countries worldwide. PloS one 16, e0246772 (2021).

38 Bushwick, S. Why COVID Vaccines Are Taking So Long to Reach You, <https://www.scientificamerican.com/article/why-covid-vaccines-are-taking-so-long-to-reach-you/> (2021).

39 Johnson & Johnson. Johnson & Johnson COVID-19 Vaccine Authorized by U.S. FDA for Emergency Use - First Single-Shot Vaccine in Fight Against Global Pandemic, <https://www.jnj.com/johnson-johnson-covid-19-vaccine-authorized-by-u-s-fda-for-emergency-usefirst-single-shot-vaccine-in-fight-against-global-pandemic> (2021).

40 Robishaw, J. D. et al. Genomic surveillance to combat COVID-19: challenges and opportunities. The Lancet Microbe (2021).

41 Cyranoski, D. Alarming COVID variants show vital role of genomic surveillance, <https://www.nature.com/articles/d41586-021-00065-4> (2021).

42 Anthes, E. Why Didn’t the U.S. Detect Omicron Cases Sooner?, <https://www.nytimes.com/2021/12/02/health/omicron-variant-genetic-surveillance.html> (2021).

43 Tindale, L. C. et al. Evidence for transmission of COVID-19 prior to symptom onset. Elife 9, e57149 (2020).

44 Cevik, M. et al. SARS-CoV-2, SARS-CoV, and MERS-CoV viral load dynamics, duration of viral shedding, and infectiousness: a systematic review and meta-analysis. The Lancet Microbe (2020).

45 Dyer, O. Covid-19: Many poor countries will see almost no vaccine next year, aid groups warn. BMJ: British Medical Journal (Online) 371 (2020).

46 Andrew, S. More than 130 countries don’t have a single Covid-19 vaccine, while 10 countries have already dispersed 75% of all vaccines, the UN says, <https://www.cnn.com/2021/02/18/world/united-nations-130-countries-no-vaccine-trnd/index.html> (2021).

47 Acharya, K. P., Ghimire, T. R. & Subramanya, S. H. Access to and equitable distribution of COVID-19 vaccine in low-income countries. npj Vaccines 6, 54, doi:10.1038/s41541-021-00323-6 (2021).

