## Supplemental Tables and Figures for "The Balancing Role of Distribution Speed against Varying Efficacy Levels of COVID-19 Vaccines under Variants"

**Supplemental Materials**

Supplemental Table 1: Infection attack rate (%) under different capacity multipliers and mutation times

| Capacity multiplier ($\lambda$) | Mutation Time = Day 10 | | | | | |  | Mutation Time = Day 15 | | | | | |
| --- | --- | --- | --- | --- | --- | --- | --- | --- | --- | --- | --- | --- | --- |
|  | $H_{i}H_{f}$ | $H_{i}M_{f}$ | $H_{i}L_{f}$ | $M_{i}M_{f}$ | $M_{i}L_{f}$ | $L_{i}L_{f}$ |  | $H_{i}H_{f}$ | $H_{i}M_{f}$ | $H_{i}L_{f}$ | $M_{i}M_{f}$ | $M_{i}L_{f}$ | $L_{i}L_{f}$ |
| 3.0 | 60.08 | 62.41 | 63.74 | 63.43 | 64.79 | 65.32 |  | 59.98 | 61.86 | 62.94 | 63.32 | 64.44 | 65.20 |
| 2.8 | 61.06 | 63.34 | 64.64 | 64.31 | 65.63 | 66.13 |  | 60.96 | 62.82 | 63.88 | 64.20 | 65.30 | 66.02 |
| 2.6 | 62.08 | 64.31 | 65.57 | 65.23 | 66.50 | 66.97 |  | 61.99 | 63.82 | 64.85 | 65.12 | 66.19 | 66.87 |
| 2.4 | 63.16 | 65.33 | 66.53 | 66.18 | 67.41 | 67.84 |  | 63.07 | 64.86 | 65.86 | 66.08 | 67.11 | 67.75 |
| 2.2 | 64.28 | 66.38 | 67.53 | 67.17 | 68.34 | 68.75 |  | 64.20 | 65.94 | 66.90 | 67.08 | 68.07 | 68.66 |
| 2.0 | 65.47 | 67.47 | 68.57 | 68.20 | 69.31 | 69.69 |  | 65.39 | 67.06 | 67.98 | 68.12 | 69.06 | 69.60 |
| 1.8 | 66.71 | 68.61 | 69.64 | 69.28 | 70.32 | 70.66 |  | 66.64 | 68.23 | 69.11 | 69.20 | 70.09 | 70.58 |
| 1.6 | 68.01 | 69.79 | 70.75 | 70.39 | 71.36 | 71.66 |  | 67.95 | 69.45 | 70.27 | 70.32 | 71.15 | 71.59 |
| 1.4 | 69.38 | 71.02 | 71.90 | 71.55 | 72.44 | 72.70 |  | 69.32 | 70.72 | 71.47 | 71.49 | 72.25 | 72.64 |
| 1.2 | 70.81 | 72.30 | 73.09 | 72.76 | 73.55 | 73.78 |  | 70.76 | 72.04 | 72.71 | 72.71 | 73.39 | 73.73 |
| 1.0 | 72.32 | 73.63 | 74.31 | 74.01 | 74.70 | 74.90 |  | 72.28 | 73.40 | 74.00 | 73.97 | 74.57 | 74.85 |

| Capacity multiplier ($\lambda$) | Mutation Time = Day 20 | | | | | |  | Mutation Time = Day 30 | | | | | |
| --- | --- | --- | --- | --- | --- | --- | --- | --- | --- | --- | --- | --- | --- |
|  | $H_{i}H_{f}$ | $H_{i}M_{f}$ | $H_{i}L_{f}$ | $M_{i}M_{f}$ | $M_{i}L_{f}$ | $L_{i}L_{f}$ |  | $H_{i}H_{f}$ | $H_{i}M_{f}$ | $H_{i}L_{f}$ | $M_{i}M_{f}$ | $M_{i}L_{f}$ | $L_{i}L_{f}$ |
| 3.0 | 59.89 | 61.39 | 62.24 | 63.22 | 64.13 | 65.10 |  | 59.75 | 60.63 | 61.14 | 63.06 | 63.62 | 64.92 |
| 2.8 | 60.87 | 62.36 | 63.21 | 64.11 | 65.00 | 65.92 |  | 60.74 | 61.63 | 62.14 | 63.95 | 64.52 | 65.75 |
| 2.6 | 61.91 | 63.38 | 64.21 | 65.03 | 65.91 | 66.77 |  | 61.78 | 62.67 | 63.18 | 64.88 | 65.44 | 66.61 |
| 2.4 | 62.99 | 64.44 | 65.25 | 66.00 | 66.85 | 67.66 |  | 62.86 | 63.76 | 64.26 | 65.86 | 66.40 | 67.51 |
| 2.2 | 64.13 | 65.54 | 66.33 | 67.00 | 67.82 | 68.57 |  | 64.01 | 64.90 | 65.39 | 66.87 | 67.40 | 68.43 |
| 2.0 | 65.32 | 66.69 | 67.45 | 68.04 | 68.83 | 69.52 |  | 65.21 | 66.08 | 66.57 | 67.92 | 68.44 | 69.39 |
| 1.8 | 66.57 | 67.89 | 68.62 | 69.13 | 69.87 | 70.51 |  | 66.47 | 67.32 | 67.80 | 69.01 | 69.51 | 70.39 |
| 1.6 | 67.89 | 69.14 | 69.82 | 70.26 | 70.96 | 71.53 |  | 67.79 | 68.62 | 69.07 | 70.15 | 70.63 | 71.42 |
| 1.4 | 69.27 | 70.44 | 71.07 | 71.44 | 72.08 | 72.58 |  | 69.18 | 69.96 | 70.39 | 71.34 | 71.78 | 72.48 |
| 1.2 | 70.72 | 71.79 | 72.37 | 72.66 | 73.24 | 73.68 |  | 70.63 | 71.37 | 71.76 | 72.57 | 72.98 | 73.59 |
| 1.0 | 72.24 | 73.20 | 73.70 | 73.93 | 74.44 | 74.81 |  | 72.16 | 72.83 | 73.19 | 73.85 | 74.22 | 74.73 |

| Capacity multiplier ($\lambda$) | Mutation Time = Day 35 | | | | | |  | Mutation Time = Day 40 | | | | | |
| --- | --- | --- | --- | --- | --- | --- | --- | --- | --- | --- | --- | --- | --- |
|  | $H_{i}H_{f}$ | $H_{i}M_{f}$ | $H_{i}L_{f}$ | $M_{i}M_{f}$ | $M_{i}L_{f}$ | $L_{i}L_{f}$ |  | $H_{i}H_{f}$ | $H_{i}M_{f}$ | $H_{i}L_{f}$ | $M_{i}M_{f}$ | $M_{i}L_{f}$ | $L_{i}L_{f}$ |
| 3.0 | 59.70 | 60.36 | 60.73 | 63.00 | 63.43 | 64.86 |  | 59.66 | 60.14 | 60.41 | 62.94 | 63.27 | 64.80 |
| 2.8 | 60.69 | 61.36 | 61.74 | 63.89 | 64.33 | 65.69 |  | 60.65 | 61.14 | 61.42 | 63.84 | 64.17 | 65.63 |
| 2.6 | 61.73 | 62.40 | 62.79 | 64.83 | 65.26 | 66.55 |  | 61.69 | 62.19 | 62.47 | 64.78 | 65.11 | 66.50 |
| 2.4 | 62.82 | 63.50 | 63.88 | 65.80 | 66.23 | 67.45 |  | 62.78 | 63.29 | 63.58 | 65.75 | 66.08 | 67.39 |
| 2.2 | 63.96 | 64.65 | 65.03 | 66.81 | 67.23 | 68.37 |  | 63.92 | 64.44 | 64.73 | 66.77 | 67.09 | 68.33 |
| 2.0 | 65.16 | 65.84 | 66.22 | 67.87 | 68.28 | 69.34 |  | 65.12 | 65.65 | 65.94 | 67.82 | 68.15 | 69.29 |
| 1.8 | 66.42 | 67.10 | 67.47 | 68.97 | 69.36 | 70.34 |  | 66.39 | 66.91 | 67.19 | 68.93 | 69.24 | 70.29 |
| 1.6 | 67.75 | 68.40 | 68.76 | 70.11 | 70.49 | 71.37 |  | 67.71 | 68.23 | 68.51 | 70.07 | 70.37 | 71.33 |
| 1.4 | 69.14 | 69.77 | 70.11 | 71.30 | 71.66 | 72.44 |  | 69.11 | 69.60 | 69.87 | 71.26 | 71.55 | 72.41 |
| 1.2 | 70.60 | 71.19 | 71.51 | 72.54 | 72.87 | 73.55 |  | 70.57 | 71.04 | 71.30 | 72.51 | 72.77 | 73.52 |
| 1.0 | 72.14 | 72.68 | 72.97 | 73.82 | 74.12 | 74.70 |  | 72.11 | 72.55 | 72.78 | 73.80 | 74.04 | 74.67 |

Supplemental Table 2: Minimum required capacity multiplier of each vaccine type under different mutation times to achieve a lower IAR than vaccine-$H_{i}H_{f}$ with the capacity multipliers of 1.0 and 1.6

| Mutation  Time | $H_{i}H_{f}$: Capacity multiplier = 1.0 | | | | |  | $H_{i}H_{f}$: Capacity multiplier = 1.6 | | | | |
| --- | --- | --- | --- | --- | --- | --- | --- | --- | --- | --- | --- |
|  | $H_{i}M_{f}$ | $H_{i}L_{f}$ | $M_{i}M_{f}$ | $M_{i}L_{f}$ | $L_{i}L_{f}$ |  | $H_{i}M_{f}$ | $H_{i}L_{f}$ | $M_{i}M_{f}$ | $M_{i}L_{f}$ | $L_{i}L_{f}$ |
| 5 | 1.24 | 1.40 | 1.28 | 1.45 | 1.48 |  | 1.97 | 2.23 | 2.04 | 2.33 | 2.37 |
| 10 | 1.2 | 1.33 | 1.28 | 1.43 | 1.48 |  | 1.91 | 2.11 | 2.04 | 2.27 | 2.37 |
| 15 | 1.17 | 1.27 | 1.28 | 1.4 | 1.47 |  | 1.85 | 2.01 | 2.04 | 2.23 | 2.36 |
| 20 | 1.14 | 1.22 | 1.27 | 1.38 | 1.47 |  | 1.81 | 1.93 | 2.03 | 2.19 | 2.35 |
| 25 | 1.12 | 1.18 | 1.27 | 1.36 | 1.47 |  | 1.77 | 1.86 | 2.03 | 2.16 | 2.35 |
| 30 | 1.1 | 1.15 | 1.27 | 1.34 | 1.46 |  | 1.73 | 1.81 | 2.03 | 2.13 | 2.34 |
| 35 | 1.08 | 1.12 | 1.27 | 1.33 | 1.46 |  | 1.7 | 1.76 | 2.03 | 2.11 | 2.34 |
| 40 | 1.06 | 1.09 | 1.27 | 1.31 | 1.46 |  | 1.68 | 1.72 | 2.03 | 2.09 | 2.34 |
